# Long-term COVID-19 symptoms in a large unselected population

**DOI:** 10.1101/2020.10.07.20208702

**Authors:** Elizabeth T. Cirulli, Kelly M. Schiabor Barrett, Stephen Riffle, Alexandre Bolze, Iva Neveux, Shaun Dabe, Joseph J. Grzymski, James T. Lu, Nicole L. Washington

## Abstract

It is increasingly recognized that SARS-CoV-2 can produce long-term complications after recovery from the acute effects of infection. Here, we report the analysis of 32 self-reported short and long-term symptoms in a general adult population cohort comprised of 357 COVID-19+ cases, 5,497 SARS-CoV-2-negative controls, and 19,095 non-tested individuals. The majority of our COVID-19+ cases are mild, with only 9 of the 357 COVID-19+ cases having been hospitalized. Our results show that 36.1% of COVID-19+ cases have symptoms lasting longer than 30 days, and 14.8% still have at least one symptom after 90 days. These numbers are higher for COVID-19+ cases who were initially more ill, 44.9% at 30 days and 20.8% at 90 days, but even for very mild and initially asymptomatic cases, 21.3% have complications persist for 30 days or longer. In contrast, only 8.4% of participants from the general untested population develop new symptoms lasting longer than 30 days due to any illness during the same study period. The long-term symptoms most enriched in those with COVID-19 are anosmia, ageusia, difficulty concentrating, dyspnea, memory loss, confusion, chest pain, and pain with deep breaths. In addition to individuals who are initially more sick having more long-term symptoms, we additionally observe that individuals who have an initial symptom of dyspnea are significantly more likely to develop long-term symptoms. Importantly, our study finds that the overall level of illness is an important variable to account for when assessing the statistical significance of symptoms that are associated with COVID-19. Our study provides a baseline from which to understand the frequency of COVID-19 long-term symptoms at the population level and demonstrates that, although those most likely to develop long-term COVID-19 complications are those who initially have more severe illness, even those with mild or asymptomatic courses of infection are at increased risk of long-term complications.

## Introduction

As time has passed since the beginning of the SARS-CoV-2 pandemic, it has become increasingly apparent that a subset of infected individuals develop long-term complications. Physicians continue to report their observations of case studies in patients who seem unable to recover from COVID-19, including long-term symptoms such as shortness of breath, weakness, brain fog, and fatigue^1–5^. Longitudinal studies of hospitalized COVID-19 patients have found that 74-88% had symptoms lasting longer than 50-80 days, most commonly fatigue and dyspnea (difficulty breathing)^6,7^. Longitudinal followup studies with quantitative phenotyping have started to elucidate the long-term sequelae in severe cases. For example, serial CT scans in hospitalized COVID-19 patients showed progression and continuation of lung abnormalities up to 24 days from onset of illness^8^. Similarly, a study using cardiac magnetic resonance (CMR) imaging in COVID-19 patients showed that 78% had abnormal findings 2-3 months after the onset of COVID-19^9^, and another study found that up to 40% of COVID-19 patients presented with pericarditis or myocarditis >70 days post-infection^10^. New and permanent loss of kidney function has also been reported in COVID-19 patients ^11^.

While there have been many case reports of the most severe forms of COVID-19, the majority of the population does not develop symptoms severe enough to require hospitalization. It is known that there is a spectrum of initial response to infection, ranging from asymptomatic individuals, to those with only a few minor symptoms who stay home, and up to those who are hospitalized. For many people throughout this spectrum, the symptoms linger, and new symptoms may arise well after the initial onset. There is an ongoing grassroots effort among these COVID-19 “long-haul” patients and their advocates to collect and share the details of long-term symptoms^12,13^, which together have uncovered a high incidence of long-term fatigue, dyspnea, difficulty concentrating, and other symptoms. However, a scientifically rigorous study of the long-term effects of COVID-19 from the general population has not yet been reported.

Here, we characterize the frequency, duration, and other properties of long-term COVID-19 symptoms by reporting the results of a prospective research study of the general population represented by participants in the Helix DNA Discovery Project^14^ and the Healthy Nevada Project^15^. We administered periodic longitudinal questionnaires to collect self-reported phenotypes to capture the occurrence and duration of COVID-19 symptoms, COVID-19 infection status, and other long-term outcomes in the general population, regardless of history of COVID-19 infection or test. Documenting the diversity of phenotypic presentation and long-term infection effects will enable the research and clinical communities to move beyond anecdotes^16^ to better understand the lasting health burden of SARS-CoV-2 in the overall population.

## Results

We received results from an online survey from 15,722 Helix DNA Discovery Project participants and 6,812 Healthy Nevada Project participants (Table 1)^14,15^. These are unselected Helix customers and patients in the Renown Health System who chose to consent to participate in research projects and respond to our survey. The survey takes approximately 10 minutes to complete and can be found in the supplement. The participants in this cohort are aged 18 to 89+, 64.1% are female, and 83.8% are of European ancestry (Table 1).

**Table 1.** Study and cohort information.

|  |  |
| --- | --- |
| Sample size | 21,359 |
| Median age (range) | 56 (18-89+) |
| N female (%*) | 14,407 (64.1%) |
| Ancestry N (%*) |  |
| African | 462 (2.1%) |
| East Asian | 368 (1.6%) |
| European | 18,884 (83.8%) |
| Latinx | 2063 (9.2%) |
| South Asian | 144 (0.6%) |
| Other / mixed ancestry | 613 (2.7%) |
| N with COVID-19 test (%) | 5,854 (23.4%) |
| Positive (%) | 357 (6.1%) |
| Negative (%) | 5497 (93.9%) |
| N reporting ≥1 symptom (%) | 13,838 (55.4%) |
| ≥1 symptom lasting longer than 30 days (%**) | 1308 (10.0%) |
| ≥1 symptom lasting longer than 60 days (%**) | 1220 (7.3%) |
| ≥1 symptom lasting longer than 90 days (%**) | 1128 (5.7%) |

\*The total here is adjusted to remove individuals who do not have their sex and ethnicity available: this demographic information was collected separately and was not yet available for some individuals for this round of analysis. \*\*The total here is adjusted to remove individuals who did not yet have enough days since their symptoms started to qualify.

Respondents were queried about 32 different symptoms that can be indicative of COVID-19 and whether they occurred between Jan 1, 2020 and the survey date^17^. Respondents answered surveys between April 2020 and October 2020, and those who responded were asked for longitudinal updates every 4-6 weeks. Respondents were additionally queried about whether they had taken a COVID-19 test and the result. Of the 21,359 respondents, 357 reported a positive COVID-19 test, 5,497 a negative test, and 19,095 were not tested. This represented a population study as opposed to one based on patients seen at the hospital: only 9 of the 357 COVID-19+ cases (2.5%) reported having been hospitalized.

### Self-reported symptoms at illness onset

Of the respondents who reported a positive COVID-19 test, 90.8% reported having ≥1 symptom during the surveyed time period, with a median of nine symptoms reported per sick person. For negative testers, 60.0% reported ≥1 symptom and a median of five symptoms per sick person, and for the remaining untested population, 53.5% reported ≥1 symptom and a median of four symptoms per sick person during the same time period (Figure 1). Because the assessed period covered a timeframe of ten months (January - October 2020), most individuals had been sick at least once during the period and thus reported at least one symptom. Though several of the surveyed symptoms were common to seasonal flu, the common cold, and allergies, we found that nearly all symptoms were statistically significant in their association with a positive COVID-19 test, even when accounting for age, sex, comorbidities, body mass index (BMI), and ethnicity (multivariate logistic regression, p<0.001). The only symptoms that were not significantly enriched in COVID-19+ cases were rash and red, sore, itchy eyes, although the frequencies of discolored digits and blisters were too rare to accurately assess statistical significance.

**Figure 1.**
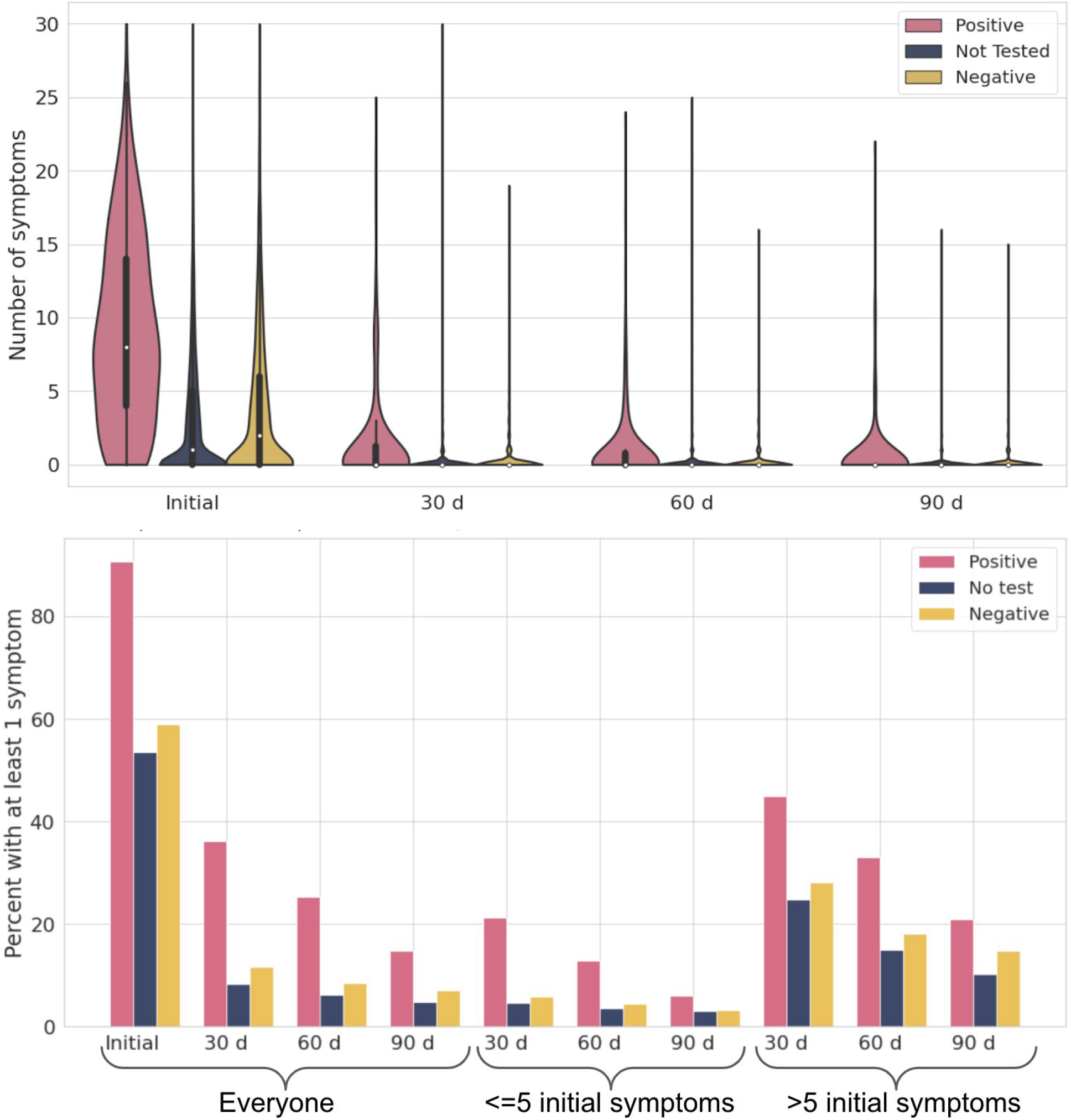
Frequency and duration of symptoms in study participants. A) Violin plot showing the number of symptoms reported in all individuals at each timepoint. B) Percent of participants with at least one symptom during the study period (initial), or at least one symptom that lasted longer than 30 days, 60 days, or 90 days. For the followup timepoints, participants are split into those who initially had 5 or fewer symptoms and those who had more (>5). Individuals whose symptoms had started less than 30, 60, or 90 days ago were excluded from their respective panels. The study period covered any illness over a ten month period (January - October 2020), and most participants reported at least one symptom occurring during that time frame.

### Accounting for overall level of illness

Although most individual symptoms assessed here were significantly enriched in COVID-19+ cases, it is also true that the total number of symptoms reported by each individual with a positive COVID-19 test was higher than the total number of symptoms reported by each person with a negative or no test, even among individuals who felt ill (Figure 1; Figure S1). In other words, if we use the number of different symptoms a person reported as a proxy for the severity of illness, then individuals with a positive COVID-19 test were generally more ill than individuals without. In fact, the total number of symptoms reported by each individual can be used to predict their COVID-19 test status (AUC = 0.76), though this method is not as accurate as employing the COVID-19-tailored symptom-based prediction model previously published by Menni et al. and confirmed in our own previous work (AUC 0.83; see Figure S2)^18,19^.

We therefore used the total number of initial symptoms reported by each person as a proxy for their severity of illness. After including this measurement as a covariate in our analyses where we assessed which symptoms were significantly enriched in COVID-19+ cases versus controls, the only symptoms that were significantly enriched in COVID-19+ cases were anosmia, ageusia, fever, fatigue, and decreased appetite, confirming previous reports about the predictive power of these symptoms^18,20^ (p<0.001; Figure 2; Table S1). However, after correcting for the total number of symptoms, many symptoms that were previously reported as associated with COVID-19, including bone or joint pain, dry cough, diarrhea, and sore muscles, were no longer significantly associated^18,20^. Additionally, we found several symptoms that were significantly depleted in positive cases after making this correction: sore throat (consistent with a previous study^20^); red, sore or itchy eyes; phlegm; cough with mucus; runny nose; and tingling somewhere in the body.

**Figure 2.**
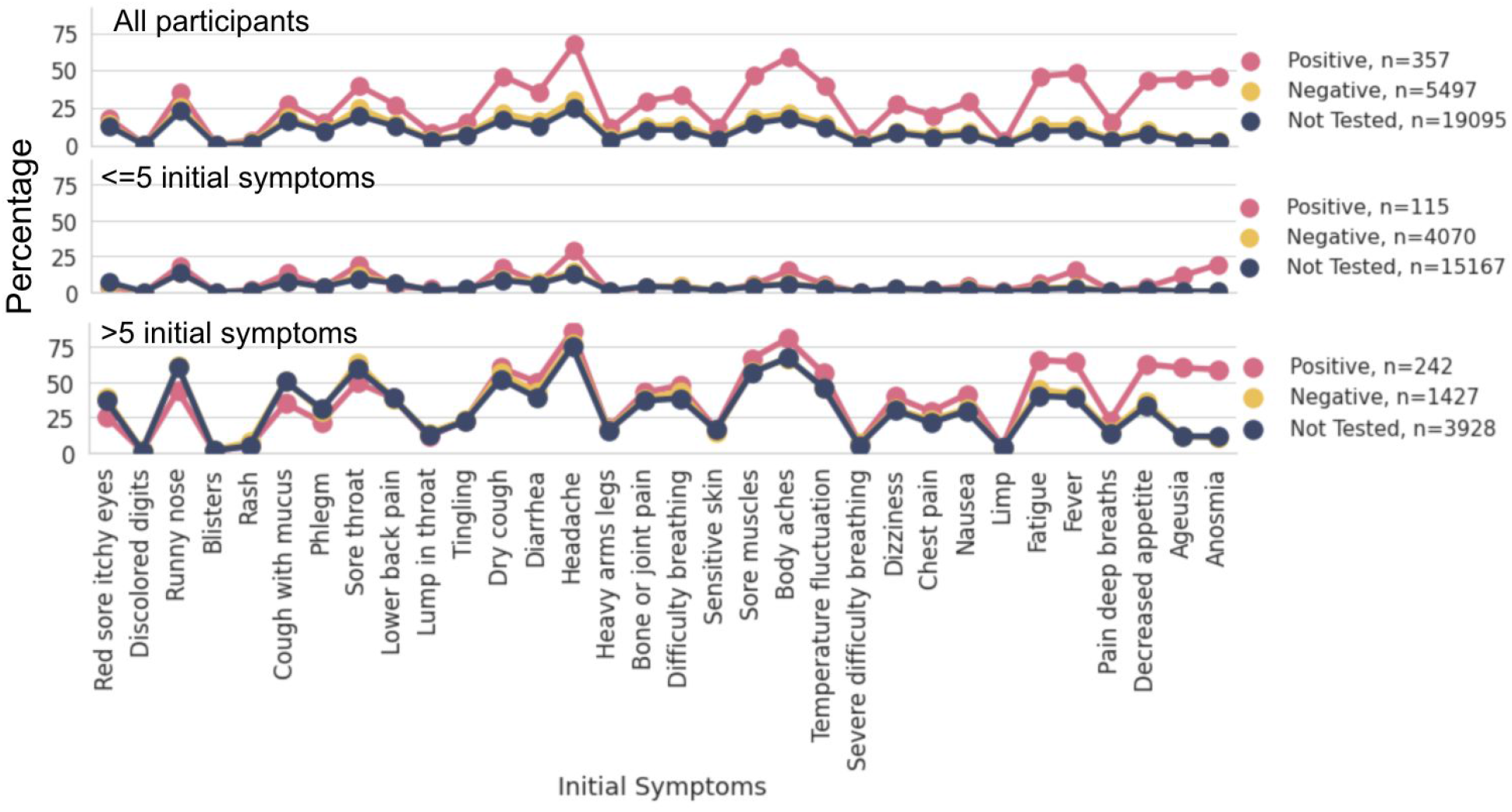
Shown is the percentage of people reporting each symptom, split into COVID-19+ cases, COVID-19-controls, and all others (Not Tested). Symptoms are ordered according to their enrichment in COVID-19+ cases vs. those with a negative test in the total sample, ordered left to right. Panel B shows only those with <=5 initial symptoms (the less ill subgroup), and Panel C shows only individuals with >5 initial symptoms (the more ill subgroup).

In addition to utilizing the total number of symptoms reported by each individual as a quantitative trait, we also stratified our comparison of symptoms to groups of those who were less ill (≤5 symptoms) and those who were more ill (>5 symptoms). This split captured the 68% of COVID-19+ cases, 26% of COVID-19-controls, and 21% of untested individuals with the most initial symptoms (Figure S1).

### Symptoms lasting longer than 30 days

We additionally asked respondents about a set of 32 long-term symptoms, defined as symptoms that lasted longer than 30 days, with initial onset occurring since the start of the pandemic. These symptoms included most of the same symptoms queried above but added symptoms that have been reported in long-haul COVID-19 patients, such as heart palpitations and memory loss. We found that after 30 days, 36.1% of our COVID-19+ cases continued to have at least 1 symptom, compared to 11.7% for those with negative tests and 8.4% for those with no test (logistic regression p=4.9×10^−22^ for a significant difference between those with a positive and negative COVID-19 test, or p=1.7×10^−5^ in a multivariate regression accounting for the total number of initial symptoms, age, sex, ethnicity, BMI, and comorbidities; Figure 3). At 60 and 90 days, these numbers were 25.3% and 14.8% for COVID-19+ cases, 8.5% and 7.0% for COVID-19-controls, and 6.3% and 4.8% for those with no tests.

**Figure 3.**
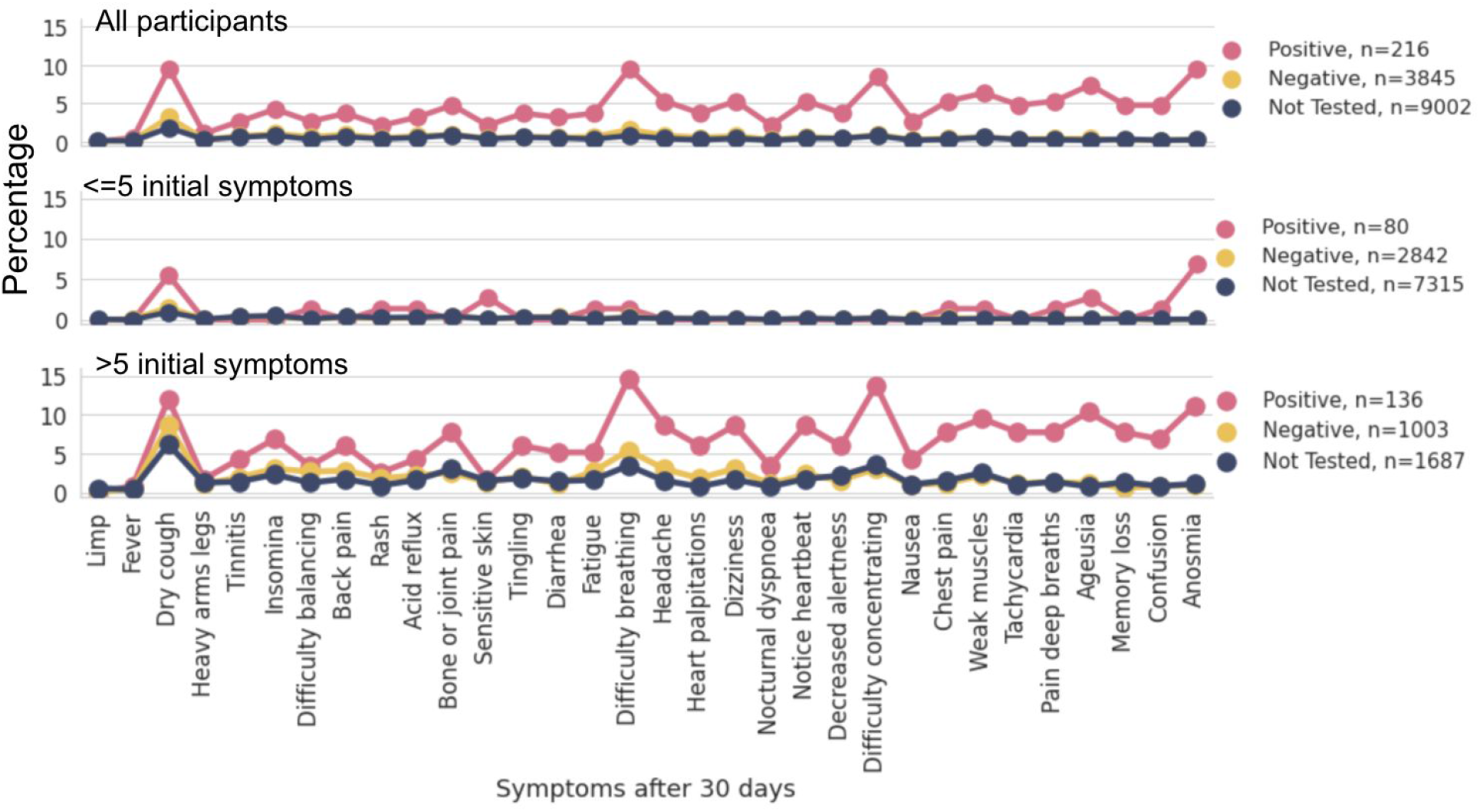
Shown is the percentage of people reporting each symptom after 30 days, split into COVID-19+ cases, COVID-19-controls, and all others (Not Tested). Symptoms are ordered according to their enrichment in those with a positive COVID-19 test vs. those with a negative test in the total sample. Panel B shows only those with <=5 initial symptoms (the less ill subgroup), and Panel C shows only individuals with >5 initial symptoms (the more ill subgroup). Symptoms after 60 and 90 days are shown in Figures S3 and S4.

The specific long-term symptoms of anosmia, ageusia, weak muscles, difficulty breathing, memory loss, confusion, difficulty concentrating, chest pain, pain with deep breaths, dry cough, decreased alertness, headache, tachycardia, bone or joint pain, heart palpitations, dizziness, nocturnal dyspnea, fatigue, tingling, sensitive skin, back pain, acid reflux, diarrhea, and insomnia were significantly enriched after 30 days in COVID-19+ cases compared to controls (p<0.001 by logistic regression; Table S2, S3, S4). However, after a multivariate regression analysis that included the initial number of symptoms in the illness as a covariate, only long-term anosmia, ageusia, memory loss, chest pain, and weak muscles remained significantly associated with COVID-19 status. With the exception of weak muscles, these symptoms remained significantly enriched in COVID-19+ cases after 60 days, at which point difficulty concentrating also became significantly enriched in COVID-19+ cases. After 90 days, all of these symptoms, plus bone / joint pain and confusion, were significantly enriched in COVID-19+ cases. Due to the relatively low numbers of people with these long-term symptoms, analysis of each individual long-term symptom was underpowered, and a larger sample size is needed to determine which of the other long-term symptoms are truly enriched in individuals with COVID-19, as well as how long they last.

As shown in Figure 1, individuals who had more initial symptoms also had more long-term symptoms, regardless of whether they were COVID-19+ cases. However, COVID-19+ cases had the highest incidence of continuing symptoms at the followup timepoints, even in the less ill category. After 30 days, 21.3% of those with initially mild cases of COVID-19 still had symptoms, though this dropped to 6.0% by 90 days. However, 20.8% of COVID-19+ cases who were initially more ill still had symptoms after 90 days. This was also true of 14.8% of individuals who were very ill during this same time period but had a negative COVID-19 test (Figure 1).

Participants were also queried about new diagnoses they had received during the study period or new procedures, such as getting a pacemaker, that they had undergone. Of the COVID-19+ cases, approximately 6% reported new diagnoses in the aftermath of COVID-19: 4% reported lung damage or scarring; 1% blood clots; 0.5% kidney damage; 0.5% heart failure; 0.5% heart damage or scarring; 0.5% myocarditis; 0.5% pulmonary embolism; and 0.5% stroke (1% reported more than one of these new diagnoses). These individuals were enriched in the subset of individuals who were initially more ill (Figure S5).

### Factors predisposing to long-term COVID-19 symptoms

We next sought to identify what factors other than seriousness of the initial illness predisposed individuals to long-term COVID-19 symptoms. Univariate logistic regression analysis of initial symptoms, comorbidities, blood types, and demographic information identified 23 factors that were at least nominally associated with whether COVID-19+ cases developed symptoms lasting longer than 30 days (uncorrected p<0.05, Figure 4, Table S5). However, the only factor that had a stronger association with symptoms after 30 days than the total number of initial symptoms (p=1.17×10^−5^) was the initial symptom of dyspnea (p=2.29×10^−6^). After including the total number of initial symptoms as a covariate in the analysis, only five factors maintained a nominal association (uncorrected p<0.05) with long-term symptoms in COVID-19+ cases: the initial symptoms of dyspnea, pain with deep breaths, and sensitive skin and blood type A were associated with increased risk; protective factors were an initial symptom of sore muscles or nausea and higher BMI. However, none of these factors remained even nominally associated with long-term symptoms after 60 and 90 days in the multivariate regression. These suggestive associations require confirmation, and the only risk factors that the data strongly support predisposing to symptoms lasting at least 30 days are dyspnea and a large number of initial symptoms.

**Figure 4.**
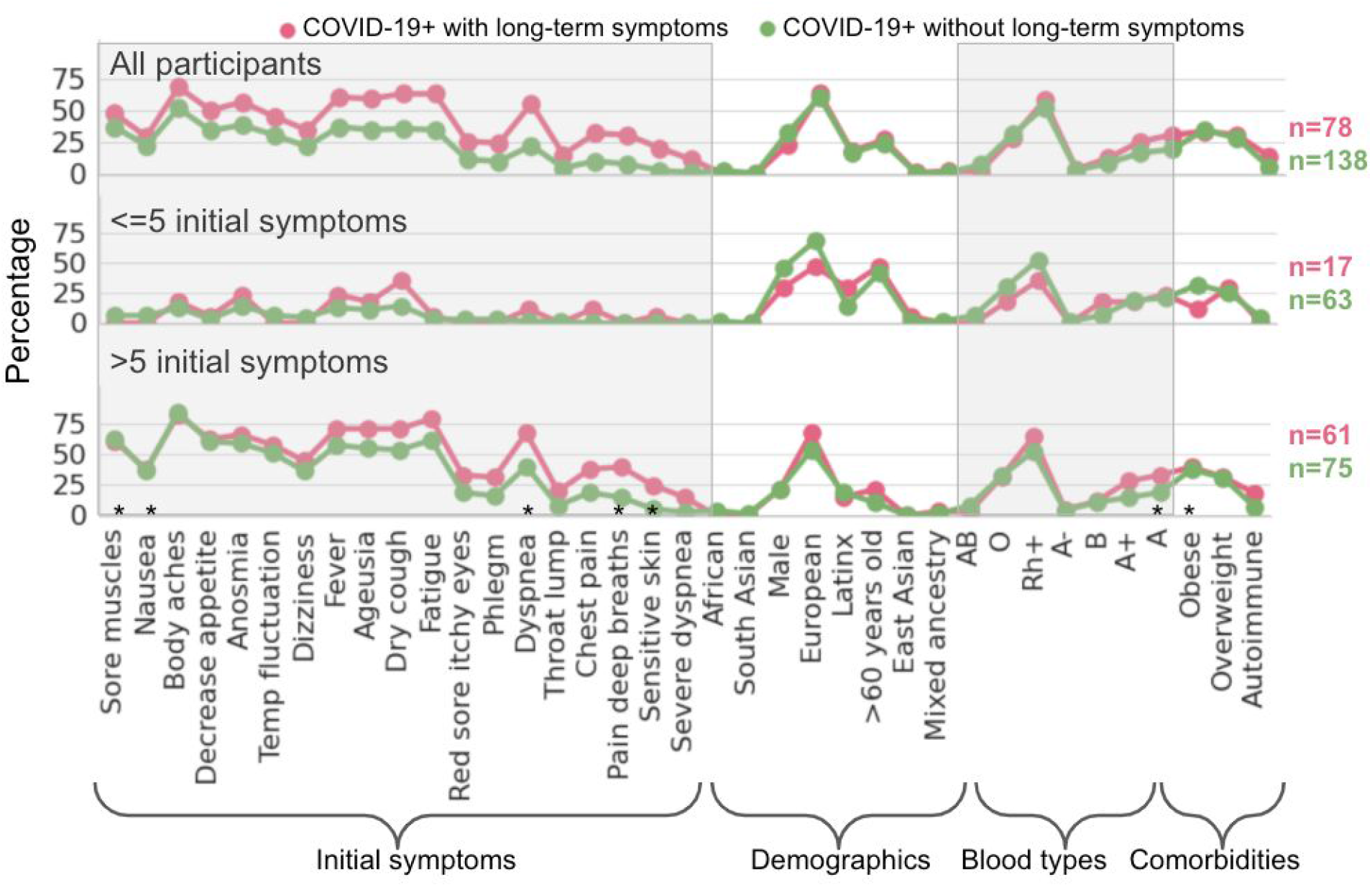
Association of different factors with long-term COVID-19 symptoms. Only COVID-19+ cases are shown, split into those who did or did not have symptoms remaining after 30 days. Shown is the percentage of people with each initial symptom, comorbidity, blood type, or demographic variable. Within categories, factors are ordered by increasing levels of enrichment in those with long-term symptoms. Panel B shows only those with <=5 initial symptoms (the less ill subgroup), and Panel C shows only individuals with >5 initial symptoms (the more ill subgroup). Most factors were nominally associated with long-term symptoms (uncorrected p<0.05); sex, demographics, and blood types beyond A were added for comparison. However, after accounting for the total number of initial symptoms, which was the strongest predictor of long-term symptoms, the only factors to maintain a nominal association (uncorrected p<0.05) were the initial symptoms of dyspnea, pain with deep breaths, sensitive skin, sore muscles, and nausea as well as blood type A and BMI (marked with *).

Although our non-European ancestry sample size is limited, we did not observe differences between ethnicities in the overall rates of long-term symptoms. The presence of pre-COVID-19 comorbidities such as diabetes and chronic lung disease was not significantly associated with long-term symptoms in this sample. However, there was a nominal association between having anxiety disorder or autoimmune/rheumatologic disorders and having long-term symptoms (Tables S5, S6, and S7). Additionally, there was no significant association between sex and the rate of long-term symptoms, though female patients were more likely to have long-term symptoms and were nominally associated with a higher frequency of symptoms at 60 days (p=0.02, Table S6). However, our sample size for positive patients with long-term information was limited (n=216), which reduced our power for discovery, and the lack of statistical significance here should not be taken to indicate a definitive lack of association.

## Discussion

Here, we report the rates of long-term symptoms consistent with COVID-19 in an unselected adult population. Importantly, our study accounts for the overall level of illness of both COVID-19+ cases and controls when assessing the statistical significance of symptoms that are associated with COVID-19. We identify that individuals who were more ill at the onset of symptoms are at higher risk of long-term symptoms. We additionally find that COVID-19+ cases who initially experienced dyspnea are at higher risk of long-term symptoms. Our population-based survey identifies a high frequency of long-term COVID-19 symptoms even among individuals who have relatively mild illness, as only nine of our participants were hospitalized.

Our study presents symptom information at the population-level, not only about COVID-19 patients but also about those who tested negative for SARS-CoV-2 as well as untested individuals from the general population. Previous studies of long-term symptoms from COVID-19 have typically focused on distinct subsets of COVID-19 patients and in particular have not included mild cases or population controls^6,12,13^. Previous studies of hospitalized COVID-19 patients found that 74-88% of COVID-19 patients who have severe initial illness still have symptoms after 50-80 days^6,7^. Another patient-led study found that >80% of those who self-recruit into a COVID-19 study on long-term symptoms still have symptoms after 50 days^12^. We likewise find a high percentage of more severely ill patients to be still affected after 60 or 90 days, but we find that the vast majority of mild COVID-19 cases have recovered after 30-60 days (Figure 1).

Some of the long-term symptoms most prevalent in previous studies of long-haul COVID-19 were fatigue, dyspnea, joint pain, chest pain, cough, anosmia, difficulty concentrating, headaches, difficulty sleeping, memory problems, and dizziness. We do not replicate a high incidence of long-term fatigue in COVID-19 patients, possibly because our patients did not represent a more severely affected, hospitalized group, and/or also because of the stringent wording in our study: “severe fatigue, such as the inability to get out of bed.” While we do see long-term COVID-19 patients with joint pain, insomnia, or cough, these rates at 30 days are not different from those in the general population of people who were ill from something other than COVID-19, which was not assessed in previous studies. We do confirm an enrichment of dyspnea, chest pain, headaches, dizziness, difficulty concentrating, memory loss, confusion, and anosmia/ageusia.

To date, there has been little research looking for factors that predispose to long-term COVID-19 symptoms. Our finding that the initial severity of the illness is correlated with long-term outcomes is in line with the higher rates of long-term symptoms found in previous studies of hospitalized patients and a previous observation that more severely ill hospitalized patients have more long-term symptoms^6,7^. Our study was underpowered to identify other factors predisposing to long-term symptoms, though we do identify a significant association between the initial symptom of dyspnea and symptoms lasting longer than 30 days.

Our method of stratifying by the initial level of illness also clarifies the short-term and long-term symptoms that were most enriched in COVID-19 patients compared to participants with other illnesses or who had symptoms from exposure to seasonal allergens or smoke from the wildfires that were prevalent in much of the study area this summer. While nearly all symptoms at onset were significantly enriched in COVID-19 cases compared to controls, only four were still significantly enriched when accounting for the level of illness: anosmia, ageusia, fever, and decreased appetite (Figure 2). In contrast, most of the symptoms that were significantly enriched in COVID-19 cases at 30 days were still enriched in the specific subset of individuals who were more sick at the onset (Figure 3), which demonstrates that memory loss, anosmia, ageusia, difficulty concentration, confusion, and bone / joint pain are all likely to be COVID-19-specific long-term symptoms.

Although the population-level survey of our study is a strength, it also limits our ability to capture the rates of long-term symptoms in the most severely ill COVID-19 patients. Only 9 of our 357 COVID-19+ cases (2.5%) were hospitalized as part of their illness. Furthermore, it is infeasible in this study to record the progress of the sickest individuals who succumbed to the illness itself. Nonetheless, our study is able to demonstrate that even individuals with relatively mild bouts of COVID-19 are more likely to have long-term symptoms than are the general public and people who became ill from other causes. We recognize that since the study data were collected by self-report via online surveys, this can introduce some error. It is also worth considering that some of our findings may show a decreased significance due to the likelihood that a significant proportion of study participants who were not tested for COVID-19 did actually have it. Further still, because COVID-19 tests can have relatively high false negative rates, some of those who received a negative test may have received it incorrectly or were at a point in their illness when SARS-CoV-2 was not detectable^21^. Continued improvements to COVID-19 tests and their availability in the general population will no doubt improve the ability for scientists to properly classify cases from controls and enhance the significance of these and other findings.

This study provides a resource for the community showing the frequency of different COVID-19 symptoms in patients over time at the population level. This baseline report opens the door for future studies of factors predisposing to long-term COVID-19 symptoms as well as genetic studies of susceptibility.

## Methods

### Participants and survey

We received online survey data from 15,722 Helix DNA Discovery Project participants and 6,812 Healthy Nevada Project participants (Table 1)^14,15^. These are unselected Helix customers and patients from Northern Nevada (Renown Health, Reno, Nevada) who consented to research involving their electronic medical records^15^. The administered surveys were designed with reference to the standard fields assembled by the COVID-19 Host Genetics Initiative and can be found in the supplement^17^. Surveys were administered at intervals of 4-6 weeks from April to October 2020. Participants who responded to any of the surveys were included, and they were not required to fill out multiple timepoints as the surveys asked about the timelines for symptoms that had already been experienced or were ongoing at the time of the survey. Questions about long-term symptoms started being administered in July. When multiple timepoints were available for a participant, their data were summarized across surveys, and they were included in each analysis only once.

As shown in the survey supplement and Tables S6, S7 and S8, the assessed comorbidities included cardiovascular disease, hypertension, asthma, chronic obstructive pulmonary disease, chronic bronchitis, cystic fibrosis, other chronic lung disease, type 1 diabetes, type 2 diabetes, sleep apnea, use of a home continuous positive airway pressure device at home at night, HIV, immunocompromised status, organ transplant, bone marrow transplant, rheumatoid arthritis, systemic lupus, erythematosus, multiple sclerosis, inflammatory bowel disease, celiac disease, other autoimmune or rheumatologic disease, liver disease, kidney disease or renal insufficiency, chronic muscle disease, depression, anxiety disorder, other mental health condition, dementia, Parkinson’s Disease, Alzheimer’s Disease, other neurological disease, balloon angioplasty or percutaneous coronary intervention, coronary artery bypass, congestive heart failure, myocardial infarction, peripheral vascular disease, stroke, arrhythmia, hepatitis, pancreatitis, pleural effusion, ascites / excess abdominal fluid, leukemia, breast cancer, prostate cancer, bladder cancer, colon and rectal cancer, endometrial cancer, kidney cancer, liver cancer, melanoma, non-Hodgkin lymphoma, pancreatic cancer, thyroid cancer, other cancer, and other chronic disease.

### Analysis

Individuals who entered contradictory information, such as marking at least one symptom but also marking “None” for symptoms, had their data excluded. Additionally, symptoms that started before 2020 were excluded. For analysis of the followup timepoints, individuals whose initial symptoms, if any, had started less than the specified number of days ago were excluded. Additionally, COVID-19-individuals were excluded from the analysis if they had received a diagnosis of COVID-19 despite their negative test.

Statistical analysis was performed using logistic regression in python. P-value thresholds were set for Bonferroni correction for multiple tests.

## Data Availability

Counts of individuals with and without symptoms in cases and controls are available in the supplement.

