## Supplementary Figures for "Long-term COVID-19 symptoms in a large unselected population"

### Supplement

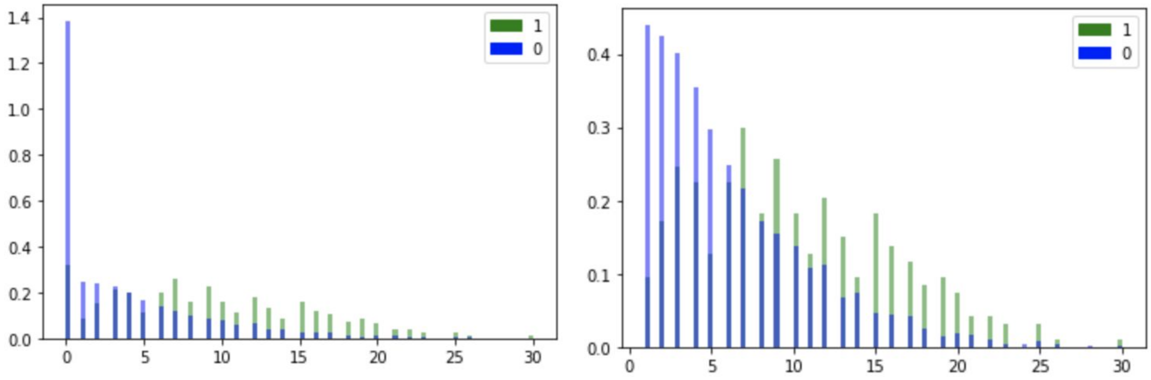


Figure S1. Overlaid histograms showing the density of people with A) 0-32 or B) 1-32 initial symptoms reported. People with a positive COVID-19 test are shown in blue, people with a negative COVID-19 test are shown in green.

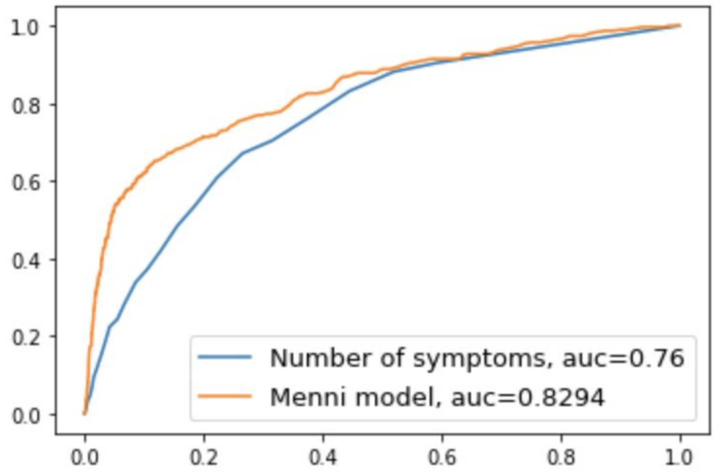


Figure S2. ROC showing performance of the model from Menni et al[18] and the number of symptoms for predicting those with a positive vs. negative COVID-19 test result in this dataset. While the Menni model clearly performs better, the total number of symptoms on its own is significantly predictive of COVID-19 test status.

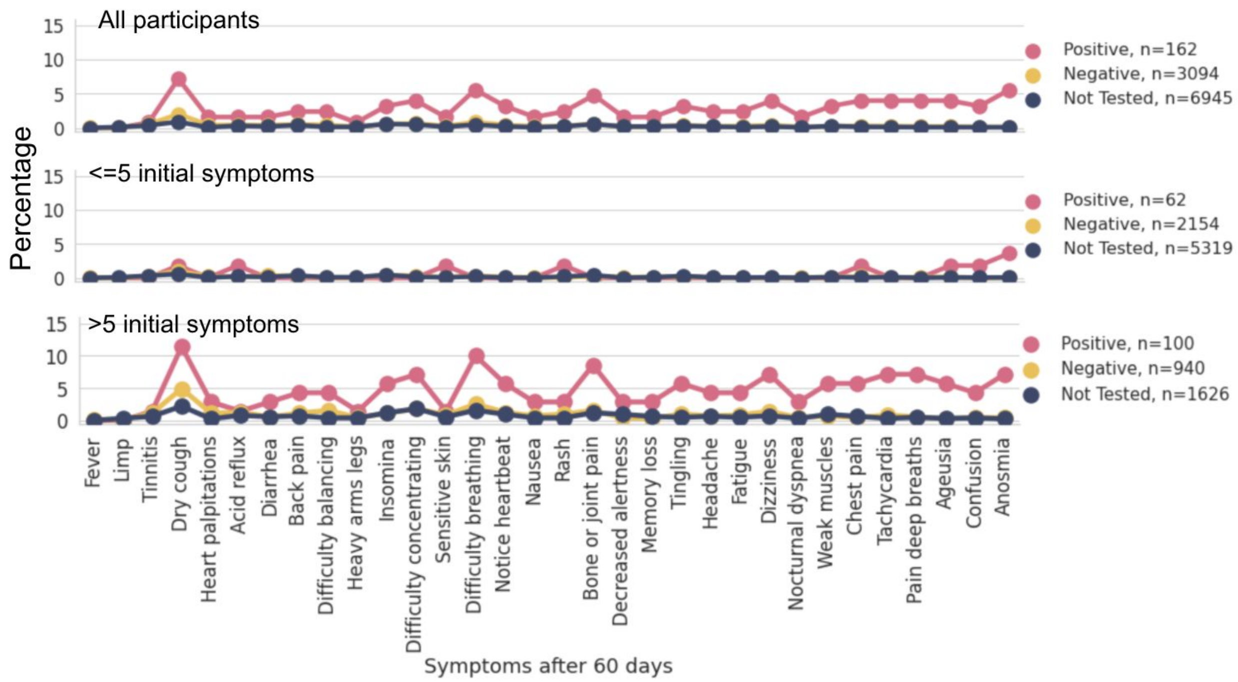


Figure S3. Shown is the percentage of people reporting each symptom after 60 days, split into individuals with a positive COVID-19 test, individuals with a negative COVID-19 test, and all others (Not Tested). Symptoms are ordered according to their enrichment in those with a positive COVID-19 test vs. those with a negative test in the total sample. Panel B shows only those with <=5 initial symptoms (the less ill subgroup), and Panel C shows only individuals with >5 initial symptoms (the more ill subgroup).

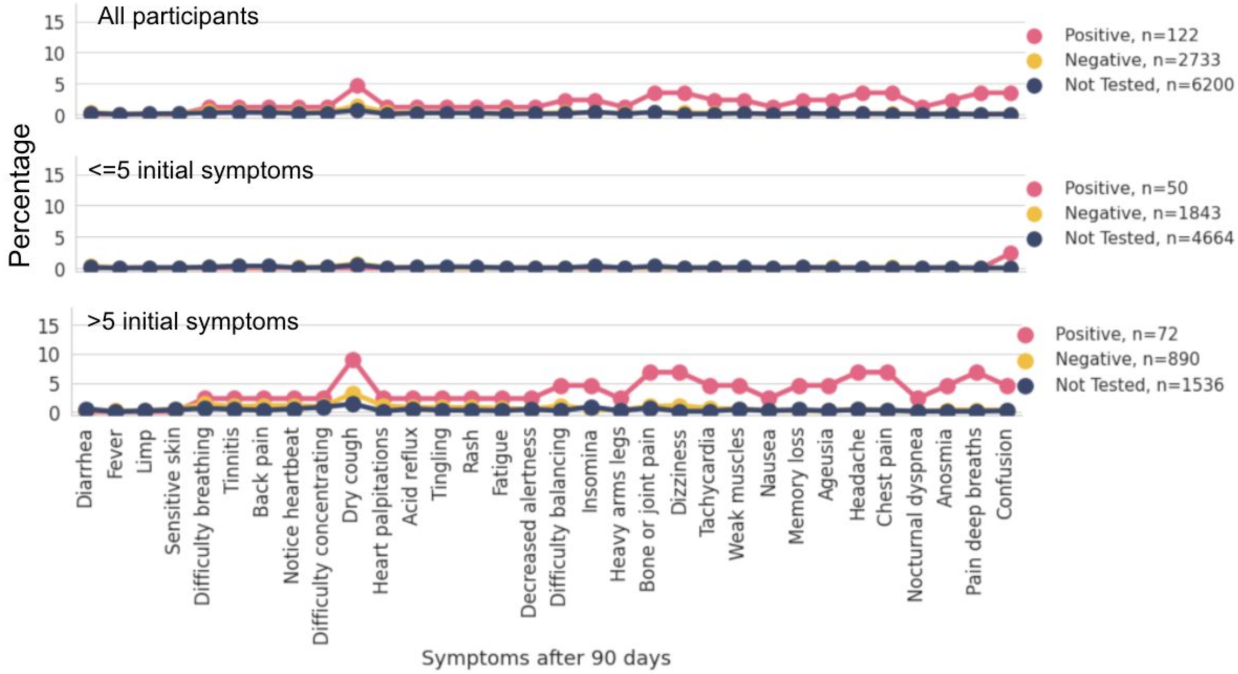


Figure S4. Shown is the percentage of people reporting each symptom after 90 days, split into individuals with a positive COVID-19 test, individuals with a negative COVID-19 test, and all others (Not Tested). Symptoms are ordered according to their enrichment in those with a positive COVID-19 test vs. those with a negative test in the total sample. Panel B shows only those with <=5 initial symptoms (the less ill subgroup), and Panel C shows only individuals with >5 initial symptoms (the more ill subgroup).

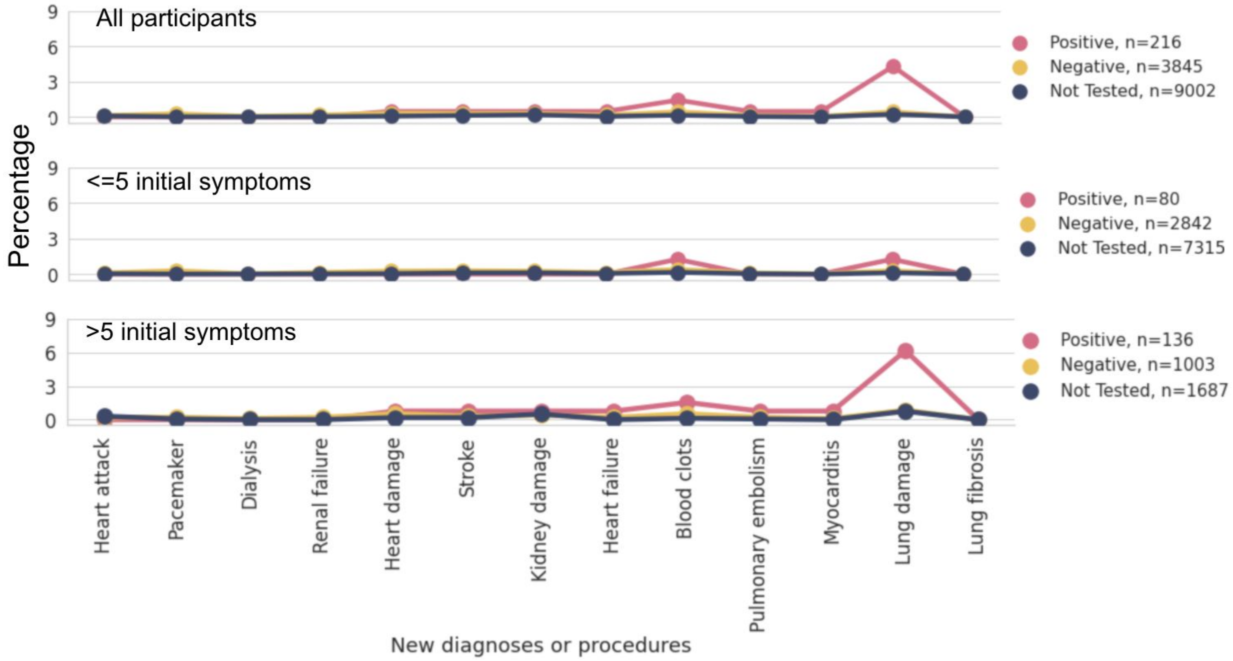


Figure S5. Shown is the percentage of people reporting new onset of a disease or a procedure, split into individuals with a positive COVID-19 test, individuals with a negative COVID-19 test, and all others (Not Tested). Panel B shows only those with <=5 initial symptoms (the less ill subgroup), and Panel C shows only individuals with >5 initial symptoms (the more ill subgroup).
